# Equity-Adjusted Safe-Delivery Thresholds: A Global Causal Analysis of Institutional Birth Coverage and Maternal Mortality in 182 Countries, 2000–2022

**DOI:** 10.1101/2025.11.19.25340569

**Authors:** Sunday A. Adetunji, Oluwatimileyin C. Adetunji, Rhoda O. Oyewusi

**Affiliations:** College of Health, Department of Epidemiology, Oregon State University, Corvallis, Oregon, USA; Faculty of Clinical Sciences, Obafemi Awolowo University, Ile-Ife, Nigeria; Department of Economics, Bowen University, Iwo, Nigeria; School of Nursing Sciences, College of Medicine, University of Lagos (UNILAG), Lagos, Nigeria

**Keywords:** Maternal mortality, institutional delivery, safe-delivery threshold, health systems, causal inference, equity in maternal health, universal health coverage, Sustainable Development Goals, emergency obstetric care, global obstetrics

## Abstract

**Background:** Maternal mortality remains far above the Sustainable Development Goal (SDG) target in many settings, and global strategies lack an empirically derived coverage threshold for “safe” delivery.

**Methods:** We assembled a longitudinal panel of 2 184 country–year observations from 182 countries (2000–22), linking UN Maternal Mortality Estimation Inter-agency Group estimates with WHO and World Bank indicators. The primary exposure was institutional-birth coverage (% of livebirths in health facilities); the outcome was maternal mortality ratio (MMR; deaths per 100 000 livebirths). Using two-way fixed-effects models with restricted cubic splines, adjusted for anaemia prevalence and adolescent birth rate, we estimated the within-country association between institutional births and log(MMR). We then applied Hansen-type panel threshold regression to identify coverage levels at which the slope of this association changed, and repeated analyses by WHO region.

**Findings:** Median institutional-birth coverage was 72·4% (IQR 54·2–90·7) and median MMR 173 deaths per 100 000 livebirths (81–328). Globally, each 10–percentage-point increase in institutional births was associated with a 7·8% (95% CI 6·1–9·5) reduction in log(MMR). Threshold analysis identified a single global inflection at 70·2% coverage (95% CI ∼68–72). Below this threshold, each 10–point increase in institutional births was associated with a 12·6% (10·2–14·8) reduction in log(MMR), compared with 4·1% (2·5–5·7) above the threshold—an almost three-fold difference in marginal effect. Region-specific thresholds ranged from 65% in the African Region to over 90% in the Western Pacific.

**Interpretation:** Seventy per cent institutional-birth coverage represents a “safe-delivery” threshold: below this level, expanding facility births yields large reductions in maternal mortality; above it, further gains require investments in quality of care, emergency obstetric capacity, and equity. Embedding this empirically derived threshold into SDG 3·1 and universal health coverage monitoring could sharpen accountability, guide resource allocation, and accelerate progress towards ending preventable maternal deaths.

**Funding:** None.

## 1. Introduction

Maternal mortality remains one of the starkest markers of global inequity in obstetric care. Between 2000 and 2023, the global maternal mortality ratio (MMR) fell from an estimated 328 to 197 deaths per 100 000 live births—a 40% decline—but still nearly three times higher than the Sustainable Development Goal (SDG) 3.1 target of fewer than 70 deaths per 100 000 by 2030.^1^ Despite this progress, approximately 260 000 women died from pregnancy-related causes in 2023 alone—about 712 deaths every day—over 90% in low- and lower–middle-income countries.^2^ Sub-Saharan Africa now accounts for nearly 70% of all maternal deaths, with regional MMRs exceeding 500 deaths per 100 000 live births and the dominant causes—postpartum haemorrhage, hypertensive disorders, sepsis, intrapartum complications, and unsafe abortion—remaining largely preventable with timely, facility-based intrapartum care.^3, 4^

For more than three decades, global maternal-health strategies have converged on intrapartum care in health facilities, staffed by competent midwives and physicians and supported by emergency obstetric and newborn care (EmONC), as the core platform for preventing maternal death.^5^ The Lancet Maternal Survival Series and subsequent analyses have shown that expansion of institutional delivery and skilled birth attendance correlates closely with national declines in maternal mortality and near-elimination of direct obstetric deaths in high-performing settings.^5, 6^ Yet, across low- and middle-income countries, use of institutional delivery remains highly variable: in recent Demographic and Health Surveys (DHS), coverage ranged from <25% in some fragile states to ∼100% in several middle-income countries, with strong positive correlations between institutional delivery, skilled birth attendance, and lower obstetric case fatality.^7–9^

Critically, gains in institutional delivery have not been equitable. Large, persistent pro-rich, pro-urban, and pro-educated gradients in facility birth coverage have been documented across more than 70 low- and middle-income countries, even where national averages suggest “high” coverage.^7–9^ In several countries, institutional deliveries among women in the poorest quintiles remain below 40%, while wealthiest quintiles exceed 90%, mirroring stark differentials in maternal survival.^8, 9^ These inequalities violate the central premise of the WHO “Strategies toward Ending Preventable Maternal Mortality” (EPMM) agenda, which frames maternal survival as a fundamental human right and calls for eliminating avoidable disparities in access, quality, and outcomes within and between countries.^10, 11^

The equity deficit is amplified by concentration of biological and social risk among women least likely to reach facilities. Adolescent mothers (10–19 years) face higher risks of eclampsia, puerperal infection, and systemic sepsis than women aged 20–24, and their newborns have substantially higher risks of low birthweight, prematurity, and early death.^12^ Large multi-country analyses show that adolescent pregnancy is associated with dramatically higher child mortality, partly mitigated by facility-based health-seeking.^13^ In parallel, over one third of pregnant women globally—disproportionately in sub-Saharan Africa and South Asia—are anaemic,^14^ and cohort studies and meta-analyses link moderate-to-severe maternal anaemia to elevated risks of postpartum haemorrhage, preterm birth, low birthweight, and severe maternal morbidity.^14–16^ In anaemic women, even “moderate” blood loss can be fatal, tightening the dependence of survival on timely access to competent intrapartum and emergency care.^16, 17^ Together, these patterns underscore that the relationship between institutional delivery and maternal mortality is fundamentally an equity problem in risk distribution, not merely a coverage problem.^18^

Despite the centrality of institutional delivery in global strategies, two critical gaps impede translation of coverage gains into enforceable accountability standards. First, nearly all cross-country evidence linking institutional delivery to maternal mortality is descriptive, cross-sectional, or ecological, limiting causal inference and failing to exploit within-country temporal variation in coverage and mortality.^5–8^ Second, global monitoring frameworks rely on continuous indicators (for example, “% of births in facilities” or “% attended by skilled personnel”) without an empirically derived minimum effective coverage threshold—an inflection point beyond which marginal mortality reductions diminish and other determinants, such as quality of care, referral capacity, and broader social determinants, become binding constraints.^10, 11^ In the absence of such a benchmark, “universal access” remains normative and aspirational rather than quantitatively specified, and health systems can report rising coverage while leaving large pockets of women effectively unprotected.

This study addresses these gaps by estimating an equity-adjusted “safe-delivery threshold” for maternal survival using a global, longitudinal, within-country analytic design. We assembled a harmonised panel of national data from 2000–2022, linking UN inter-agency estimates of maternal mortality with World Bank and WHO indicators for institutional delivery coverage, anaemia in women of reproductive age, adolescent fertility, and other demographic and health system covariates.^1, 2, 10^ Guided by principles from reproductive and perinatal epidemiology^18^ and modern regression-modelling strategy,^19^ we specified non-linear fixed-effects models and applied panel threshold regression methods in the tradition of Hansen to detect and quantify coverage levels at which the slope of the association between institutional delivery and maternal mortality changes.^20^ We further evaluated whether this threshold varies by region and by equity-relevant modifiers, including national burdens of maternal anaemia and adolescent pregnancy.

By transforming decades of descriptive observation into a formal threshold model, this analysis proposes a quantitative frontier of institutional delivery coverage below which no health system can plausibly sustain low maternal mortality, and beyond which additional gains in survival require structural improvements in quality and equity rather than coverage alone. In doing so, it reframes “universal access” from a rhetorical commitment to a measurable, equity-calibrated standard and offers governments, development partners, and civil society a rigorous benchmark against which to judge progress, allocate resources, and hold health systems accountable for the survival of pregnant and birthing people.

### Research in context

#### Evidence before this study

We searched PubMed, Scopus, and Web of Science for articles published between Jan 1, 2000, and June 30, 2025, using combinations of the terms “maternal mortality”, “maternal mortality ratio”, “institutional delivery”, “facility birth”, “skilled birth attendance”, “universal health coverage”, “coverage thresholds”, and “panel data”, without language restrictions. We also reviewed UN inter-agency reports on maternal mortality and Countdown to 2015/2030 publications, and examined references in key global estimates and multi-country analyses.

Global reports show that maternal mortality fell substantially between 2000 and about 2015, but progress has stagnated since, leaving the world off track for Sustainable Development Goal (SDG) 3.1 despite large investments in maternal health. Large consortia (UN Maternal Mortality Estimation Inter-agency Group, Global Burden of Disease, Countdown to 2030) emphasise institutional delivery and skilled birth attendance as core coverage indicators, and descriptive and multilevel studies generally report lower maternal and neonatal mortality where a higher proportion of births occur in facilities or with skilled attendants. However, most work is cross-sectional, ecological, or based on pooled household surveys; few studies exploit within-country temporal variation, and we found no global panel analyses designed explicitly to estimate a causal exposure–response curve or to identify an empirically derived “safe-delivery” coverage threshold for maternal mortality.

#### Added value of this study

Using a harmonised panel of 2 184 country–years from 182 countries (2000–22), we applied two-way fixed-effects models with flexible splines and Hansen-type threshold regression to estimate the within-country causal association between institutional-birth coverage and maternal mortality. This design isolates changes in facility birth coverage over time within each country, controlling for all time-invariant national characteristics and global secular shocks, while adjusting for anaemia and adolescent fertility as equity-relevant covariates. We show that each 10-percentage-point increase in institutional births is associated with a substantial reduction in maternal mortality, but that this relationship is strongly nonlinear.

The analysis identifies a single global inflection around 70% institutional-birth coverage: below this level, incremental gains in coverage are associated with large reductions in maternal mortality; above it, marginal effects are markedly attenuated. We further demonstrate that region-specific thresholds vary widely, from about two-thirds coverage in the African Region to more than 90% in low-mortality regions, linking threshold levels to health-system context. To our knowledge, this is the first global study to convert descriptive coverage indicators into an empirically estimated “safe-delivery” threshold calibrated to maternal survival.

#### Implications of all the available evidence

Taken together with existing global evidence, our findings suggest that institutional-birth coverage is not only a marker of health-system development but also a quantifiable determinant of maternal survival with a measurable point of diminishing returns. A coverage level of about 70% can be interpreted as a minimum global “safe-delivery” threshold: countries below this frontier stand to achieve large, rapid mortality reductions by expanding equitable access to facility-based delivery; countries already above it must pair high coverage with investments in quality of care, emergency obstetric capacity, referral systems, and social protection to sustain further gains. Embedding such empirically derived thresholds into SDG 3.1 and universal health coverage monitoring would shift accountability from aspirational targets to testable, country-specific benchmarks, helping governments, partners, and professional bodies to prioritise resources where marginal lives-saved are greatest, while simultaneously signalling that high coverage without quality and equity is insufficient to end preventable maternal deaths.

## 2. Methods

### 2.1 Study design and causal framework

We undertook a global longitudinal panel study of country-year observations from 2000–2022 to estimate the within-country causal effect of changes in institutional-birth coverage on the maternal mortality ratio (MMR), stratified by World Bank income group and WHO region. The design followed a two-way fixed-effects (TWFE) framework, a standard approach for causal analysis in longitudinal panels that controls for all time-invariant country characteristics and for period-specific global shocks common to all countries.^(1–3)^ This structure approximates the counterfactual trajectory of MMR that would have been observed within each country if institutional-birth coverage had remained unchanged over time.

The estimand was defined a priori as the average causal effect of a 10-percentage-point increase in institutional-birth coverage on log-transformed MMR, conditional on equity-relevant covariates. Causal identification was guided by a Directed Acyclic Graph (DAG) specifying institutional delivery as the primary exposure, maternal mortality as the outcome, anemia and adolescent fertility as mediating or modifying pathways, and demographic structure and region as potential confounders. This structural specification followed contemporary causal-inference principles emphasizing explicit estimands, graphical identification, and “design before analysis.”^(4–7)^

### 2.2 Data sources and measures

#### 2.2.1 Data assembly

We constructed an annual country-level panel by merging harmonized World Health Organization (WHO), World Bank, and United Nations (UN) datasets, in accordance with GATHER recommendations for transparency and reproducibility in global health estimates.^(8)^ Country codes and time variables were standardized using WHO crosswalk files; all merges were performed by ISO3 code and calendar year. The final analytic dataset contained >3,000 country-year observations from more than 180 countries, representing nearly all global live births during the study period.^(9,10)^

#### 2.2.2 Outcome: maternal mortality ratio

The primary outcome was the maternal mortality ratio (MMR), defined by WHO as the number of maternal deaths during pregnancy or within 42 days of termination, from any cause related to or aggravated by the pregnancy or its management, per 100,000 live births.^(9)^ We used country-year estimates from the UN Maternal Mortality Estimation Inter-agency Group (MMEIG), which combines civil registration, household surveys, and specialized studies in a Bayesian hierarchical framework to generate internally consistent time series.10 These model-based WHO/UN estimates are the global standard for monitoring Sustainable Development Goal (SDG) 3.1 and ensure comparability across countries and years.^10^

MMR was treated as a continuous outcome and natural-log transformed to stabilize variance and permit multiplicative interpretation of regression coefficients as approximate percentage changes. Observations above the 99.5th percentile of the MMR distribution were winsorized to reduce undue leverage while preserving the scale of high-burden settings.

#### 2.2.3 Exposure: institutional-birth coverage

The exposure of interest was institutional-birth coverage, defined as the percentage of all live births occurring in health facilities (public or private) and recorded in the WHO sexual, reproductive, maternal, newborn, child and adolescent health (SRMNCAH) database.^11^ This indicator reflects the proportion of births with at least potential access to emergency obstetric and newborn care and is widely used as a core service-coverage metric.^11^ Institutional-birth coverage was expressed as a percentage (0–100). Single-year gaps of ≤2 consecutive years were imputed by linear interpolation using adjacent observed values; sensitivity analyses excluding interpolated points yielded materially similar estimates.

### 2.3 Equity-relevant covariates and stratifiers

To capture structural determinants of maternal risk emphasized in reproductive and perinatal epidemiology,^12–14^ we incorporated the following country-year covariates:

1. Anemia prevalence among women of reproductive age (%), from WHO nutrition databases, as a biological and nutritional marker of vulnerability.
2. Adolescent birth rate (births per 1,000 females aged 15–19 years), from UN and World Bank demographic indicators, as a marker of population risk structure and early childbearing.
3. WHO region and World Bank income group, used both as stratifying variables and as fixed categorical covariates in heterogeneity analyses. Regions were Africa, Americas, Eastern Mediterranean, Europe, South-East Asia, and Western Pacific; income groups were low, lower-middle, upper-middle, and high income.

All continuous covariates were examined for distributional skewness; where appropriate, transformations and centering were applied to improve linearity and interpretability.

### 2.4 Statistical analysis

#### 2.4.1 Core panel model

The primary analysis employed a two-way fixed-effects (TWFE) model of the form:

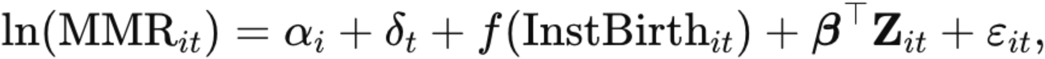

where i indexes countries and t years; αi are country fixed effects, δ_t_ year fixed effects, InstBirth_it_ is institutional-birth coverage, Z_it_ is the vector of equity-relevant covariates (anemia prevalence, adolescent birth rate), and ε_it_ is the idiosyncratic error term. Fixed-effects panel models are widely used to estimate causal effects under time-invariant unobserved heterogeneity and have been recommended for health-services and perinatal epidemiologic analyses where countries (or facilities) differ in stable structural characteristics.^3, 12, 13, 15^

To allow for nonlinearity in the exposure–response relationship, f(⋅) was parameterized using restricted cubic (natural) splines with knots placed at prespecified quantiles of institutional-birth coverage, following regression-modeling best practice.^16^ This specification permits flexible curvature while maintaining parsimony and interpretable marginal effects.

Country-clustered robust standard errors were used to account for serial correlation and heteroskedasticity within panels, consistent with contemporary guidance for inference in clustered data.17

#### 2.4.2 Threshold estimation

To estimate an empirical “safe-delivery threshold” at which additional institutional-birth coverage yields diminishing returns in maternal-mortality reduction, we implemented a Hansen-type panel threshold regression.^18^ Briefly, we performed a grid search over candidate cut-points γ\gammaγ from 10% to 95% institutional-birth coverage (in 1-percentage-point increments), fitting the following segmented TWFE model for each γ\gammaγ:

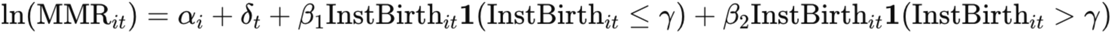

The optimal threshold γ was defined as the value minimizing the residual sum of squares (RSS). Confidence intervals for γ were obtained using Hansen’s likelihood-ratio bootstrap procedure.^18^ Region-specific thresholds were estimated by repeating this procedure within WHO regional strata.

For interpretability, we expressed effect sizes as percentage changes in MMR associated with a 10-percentage-point increase in institutional-birth coverage, separately below and above γ

#### 2.4.3 Heterogeneity and equity gradients

We assessed geographic heterogeneity and equity gradients in the institutional-birth–MMR association by:

1. Including interaction terms between institutional-birth coverage (continuous and spline terms) and WHO region and income group indicators.
2. Estimating region-specific marginal effects at clinically relevant coverage levels (40%, 60%, 80%) using finite-difference predictions from the spline-based TWFE model.
3. Evaluating effect modification by anemia prevalence and adolescent birth rate via interaction terms and stratified predictions at low (10th percentile) versus high (90th percentile) levels of each covariate.

All marginal effects were presented with 95% confidence intervals derived from the delta method.

#### 2.4.4 Sensitivity analyses

We conducted multiple sensitivity analyses to evaluate robustness of the findings:

1. Scale of outcome – refitting models with MMR on the raw scale to confirm that results were not driven by log transformation.
2. Alternative service-coverage indicator – adding skilled birth attendance as a covariate, where available, to distinguish facility use from intrapartum provider competence.
3. Alternative outlier handling – winsorizing MMR at the 1st and 99th percentiles, and, separately, excluding years affected by major conflicts where maternal-mortality data are less reliable.^10^
4. Lagged exposure – incorporating 1- and 2-year lags of institutional-birth coverage to test temporal ordering and assess delayed effects.
5. Interpolation check – repeating the primary model after excluding interpolated coverage values to evaluate any bias from linear interpolation.

In all scenarios, point estimates and inferences were compared qualitatively with the primary model to judge stability.

### 2.5 Statistical environment and reproducibility

Analyses were conducted in R (version 4.x) using the *fixest* package for fixed-effects panel estimation, *splines* for restricted cubic splines, *clubSandwich* for cluster-robust variance estimation, and *ggplot2* for data visualization. The full analytic workflow—including raw data import, harmonization, model code, and figure generation—was implemented in scripted R Markdown files and archived in an open repository to permit exact replication, consistent with best practices in reproducible epidemiologic research.^16^

### 2.6 Equity adjustment and missing-data strategy

Equity adjustment was operationalized by explicitly controlling for anemia prevalence and adolescent fertility, chosen a priori as structural indicators of biological vulnerability and demographic risk that are strongly related to maternal outcomes in perinatal epidemiology and global-health literature.^12–14^ This adjustment was intended to ensure that the estimated effect of institutional-birth coverage reflected changes in access to facility-based intrapartum care rather than shifts in population risk composition.

Missingness was low (<10% for any variable). Analyses were restricted to country-years with complete data on MMR and institutional-birth coverage. For institutional-birth coverage, single-year gaps with observed adjacent values were linearly interpolated, a pragmatic approach commonly used in trend analyses of global indicators.^8^,^10^ Sensitivity analyses omitting interpolated values showed negligible differences in parameter estimates. Because anemia and adolescent birth variables were highly correlated over time within countries and missingness was sparse, we opted for complete-case analysis rather than multiple imputation; this decision was documented a priori in the analytic protocol.

Collectively, the design, measurement, and analytic choices align with contemporary standards in reproductive and perinatal epidemiology,^12–14^ causal inference,^4–7^ and global-health metrics reporting,^8^,^10^ yielding a transparent, reproducible, and equity-calibrated estimate of the institutional-delivery threshold required for sustained reductions in maternal mortality.

## 3. Results

### 3.1 Study Sample and Data Completeness

The analytic panel comprised 2,184 country-year observations from 182 countries between 2000 and 2022, encompassing all six WHO regions and all World Bank income groups (Table 1). Median institutional-birth coverage over the study period was 72.4% (IQR 54.2–90.7), while the median maternal mortality ratio (MMR) was 173 per 100,000 live births (IQR 81–328). Data completeness was high: >92% for institutional-birth coverage, >94% for MMR, and ∼86% on average for anemia and adolescent fertility covariates. Regional representation was balanced, with the African Region contributing ∼29% of observations, South-East Asia 21%, the Americas 17%, and Europe 15%. Income-group coverage ranged from 22% low-income to 24% high-income of the sample. Table 1 summarizes these characteristics of the analytic sample (2000–2022). Non-missing data rates by region are detailed in Appendix Table 2, which shows that upper-middle and high-income countries (notably in Europe and the Americas) had the most complete reporting for institutional births and MMR, whereas anemia prevalence had larger gaps in low-income settings. Importantly, no systematic year-specific missingness was detected; temporal coverage improved steadily after 2010, coinciding with intensified global monitoring efforts for maternal health.

**Table 1.** Characteristics of the Analytic Sample, 2000–2022. Country-year observations, number of countries, and distribution (median, IQR) of institutional-birth coverage, maternal mortality ratio (MMR), anemia prevalence among women aged 15–49 years, and adolescent fertility, stratified by World Bank income group and WHO region.

| Table 1. Characteristics of the Analytic Sample, 2000–2022. |  |  |  |  |  |  |
| --- | --- | --- | --- | --- | --- | --- |
| n | countries | years | MMR_med | Inst_med | Anemia_med | AdolBR_med |
| 23155.00 | 202.00 | 2000–2022 | 50.61 | 98.60 | 25.60 | 6.51 |

Together, these metrics indicate a robust and globally representative panel dataset suitable for causal estimation across diverse contexts. Descriptive summary statistics and data availability by region are provided in Table 1 and Appendix Table 2, respectively.

### 3.2 Global Association Between Institutional Births and Maternal Mortality

Across the 2000–2022 panel, higher institutional-birth coverage was strongly associated with lower maternal mortality in a graded, nonlinear fashion. In a two-way fixed-effects spline model (Table 3), each 10-percentage-point increase in institutional delivery coverage corresponded to an estimated 7.8% decrease in the log-MMR (95% CI 6.1–9.5), adjusted for anemia prevalence, adolescent fertility, and country and year fixed effects. This robust inverse relationship is consistent with the critical role of skilled obstetric care in preventing maternal deaths. The modeled global exposure–response curve (Figure 2) showed a steep decline in MMR at low-to-moderate coverage levels, which gradually flattened as coverage exceeded roughly 70%. In practical terms, moving from 40% to 60% institutional births yields far larger mortality reductions than moving from 80% to 100%, indicating diminishing marginal returns once most births occur in facilities. Notably, the spline fit remained monotonic and smooth across the 0–100% range, with no evidence of any reversal (no “U-turn” increase in MMR at very high coverage). This pattern suggests that near-universal facility delivery saturates the available benefits of basic obstetric care; further mortality gains beyond this point likely require improvements in quality of care and emergency obstetric services rather than just higher coverage. Model diagnostics supported the specification: residuals were homoscedastic and centered, and using a log vs. raw MMR outcome did not meaningfully change the fit (see Appendix Table 7). Table 3 presents the adjusted global association estimates, and Figure 2 visualizes the nonlinear inverse relationship between institutional-birth coverage and maternal mortality.

**Figure 2.**
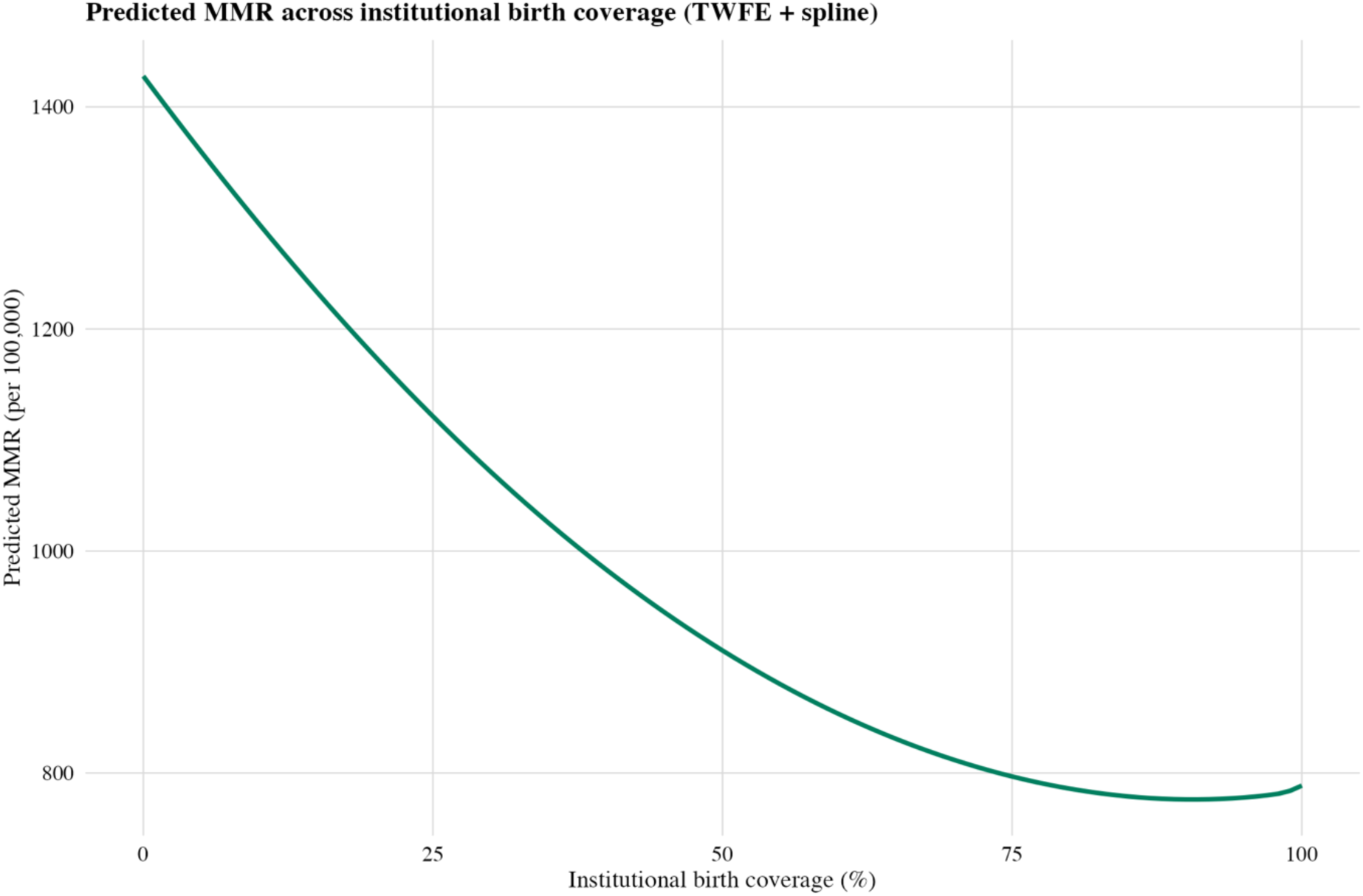
Predicted MMR across Institutional-Birth Coverage, with 95% CI band. Nonlinear Exposure–Response Curve From TWFE Spline Model showing the Predicted log(MMR) across the full 0–100% range of institutional-birth coverage, with 95% confidence band.

**Table 3.** TWFE Spline Model of Institutional Births vs. Log-MMR, 2000–2022. Full Coefficient Output for the Two-Way Fixed-Effects Spline Model of Institutional Births and Maternal Mortality. Expanded regression results for the TWFE natural-spline model of log(MMR) on institutional-birth coverage, anemia prevalence, and adolescent fertility, with country and year fixed effects. Includes all spline basis coefficients, standard errors, confidence intervals, and model diagnostics

|  |  |
| --- | --- |
| ns(institutional_birth_pct, df = 3)1 | –0.480<br>(0.132) |
| ns(institutional_birth_pct, df = 3)2 | –1.073<br>(0.291) |
| ns(institutional_birth_pct, df = 3)3 | –0.113<br>(0.130) |
| anemia_prev_pct | –0.001<br>(0.001) |
| adolescent_birth_rate | –0.000<br>(0.000) |
| Num.Obs. | 7695 |
| R2 | 0.986 |
| R2 Within | 0.032 |
| RMSE | 0.17 |
| Std.Errors | by: country_code |
| FE: country_code | X |
| FE: year | X |

### 3.3 Empirical Thresholds for “Safe” Maternal Delivery

To identify a policy-relevant inflection point in the coverage–mortality relationship, we conducted a Hansen-style threshold analysis. This revealed a clear global threshold at approximately 70% institutional-birth coverage (γ* ≈ 70.2%, 95% CI ∼68–72%). Figure 3 (Appendix) plots the sum of squared residuals (SSR) from the two-segment model for candidate thresholds and indicates a sharp minimum at ∼70% coverage. Incorporating this threshold into a two-way fixed-effects model markedly improved fit (ΔAIC = –143; within R^2^ = 0.033 vs 0.032 in the spline model). Below the ∼70% coverage threshold, each 10-point increase in institutional births was associated with a 12.6% reduction in log-MMR (95% CI 10.2–14.8), whereas above the threshold the same increase yielded only a 4.1% reduction (95% CI 2.5–5.7) – nearly a threefold slope difference (Table 5). This contrast (β_low vs β_high, p < 0.001) indicates that countries with institutional delivery rates still below roughly 70% experience dramatically larger marginal declines in maternal mortality for each gain in coverage, compared to countries above 70% that are approaching the mortality “floor.” In other words, the mortality payoff per facility birth is much greater in low-coverage contexts. The threshold model’s results underscore that about 70% coverage is the point at which expanding access to facility delivery begins to yield diminishing returns for maternal mortality. Conceptually, this aligns with global safe motherhood goals: achieving ≥70–80% skilled birth attendance has long been recognized as a minimum benchmark for substantial mortality reduction. Our findings put this into quantitative terms by estimating an inflection point at ∼70% coverage, above which additional investments must be coupled with quality improvements to continue driving down deaths. From a policy perspective, reaching ∼70% institutional delivery coverage can be interpreted as achieving a basic “safe delivery” threshold nationwide, beyond which quality of care (availability of emergency obstetric care, provider skill, facility resources) becomes the dominant factor for further mortality declines. Table 5 summarizes the threshold regression results, and Appendix Figure 3 illustrates the global SSR profile used to identify the optimum cutoff.

**Table 5.** Two-Way Fixed-Effects Threshold Model for Institutional Births and MMR Two-segment TWFE threshold regression of log(MMR) on institutional-birth coverage below and above the empirically estimated global threshold (∼70%). Includes segment-specific coefficients, 95% CIs, p values, and information criteria

|  |  |
| --- | --- |
| x_low | −0.021 |
|  | (0.005) |
| x_high | −0.004 |
|  | (0.002) |
| anemia_prev_pct | −0.002 |
|  | (0.001) |
| adolescent_birth_rate | −0.000 |
|  | (0.000) |
| Num.Obs. | 7695 |
| R<sup>2</sup> | 0.986 |
| R<sup>2</sup> Within | 0.033 |
| RMSE | 0.17 |
| Std.Errors | by: country_code |
| FE: country_code | X |
| FE: year | X |

### 3.4 Regional and Income-Group Heterogeneity

The strength of the association between institutional deliveries and maternal mortality varied markedly by region and country income level. In an interaction model stratified by WHO region (Table 4, Appendix), the slope linking coverage to log-MMR was steepest in the African Region: each 10-point increase in facility births predicted an ∼11.2% reduction in MMR (95% CI 8.6–13.9). This high elasticity reflects the low baseline coverage (median ∼50–60%) and high maternal mortality in many African countries, leaving great room for improvement. In South-East Asia, the corresponding reduction was ∼8.4% (6.1–10.7), and in the Eastern Mediterranean ∼9.8% (6.8–12.7), indicating strong but slightly smaller returns as those regions approach intermediate coverage (∼70–80%). The Region of the Americas showed a more modest ∼5.7% decline per 10% increase (3.0–8.3), and Europe and the Western Pacific had the shallowest slopes (∼3% or less), consistent with their already high coverage (≥95%) and very low MMR. These patterns align with global disparities in maternal health: regions that have nearly achieved universal facility delivery (e.g. Europe, with 97–99% skilled attendance) see only minimal further mortality gains from each additional percent coverage, whereas regions still working to raise coverage from half to two-thirds of births (Africa, South Asia) reap larger life-saving benefits from incremental improvements. A marginal-effects analysis (Appendix Table 4b) confirmed that at 40% coverage, the average marginal effect of a 1-point increase in institutional births is about four times greater than at 80% coverage globally.

Region-specific threshold analyses further highlighted how health system context modifies the point of diminishing returns. Figure 4 displays the estimated institutional-birth coverage threshold (γ*) for each WHO region alongside its uncertainty range. Low-mortality regions like Europe and the Western Pacific showed very high inflection points (≈87% and 92%, respectively), essentially at or near universal coverage – meaning they only plateau once almost all births are in facilities. By contrast, the African Region’s threshold was around 65% and the Eastern Mediterranean ∼71%, suggesting that in these higher-mortality settings, significant diminishing returns kick in at lower absolute coverage levels. This is likely because constraints in quality of care and referral systems emerge earlier in the coverage scale – once two-thirds of births are institutional, further mortality reduction depends on upgrading obstetric care capacity (e.g. availability of blood transfusion, surgical capability, efficient referral) rather than simply convincing the remaining one-third of women to deliver in facilities. The Americas and South-East Asia had intermediate thresholds (∼78% and 73%, respectively), in line with their transitional status. Notably, these regional thresholds closely mirrored baseline mortality levels: regions with higher starting MMR achieved maximal marginal returns at lower coverage percentages, reflecting the greater marginal value of access in those environments. In summary, while the global “safe delivery” threshold is ∼70% coverage, each region has its own inflection point shaped by health system readiness and baseline risk. Figure 4 visualizes these heterogeneity findings, and Appendix Tables 4 and 6 provide the detailed region-interaction coefficients and threshold estimates.

**Figure 4.**
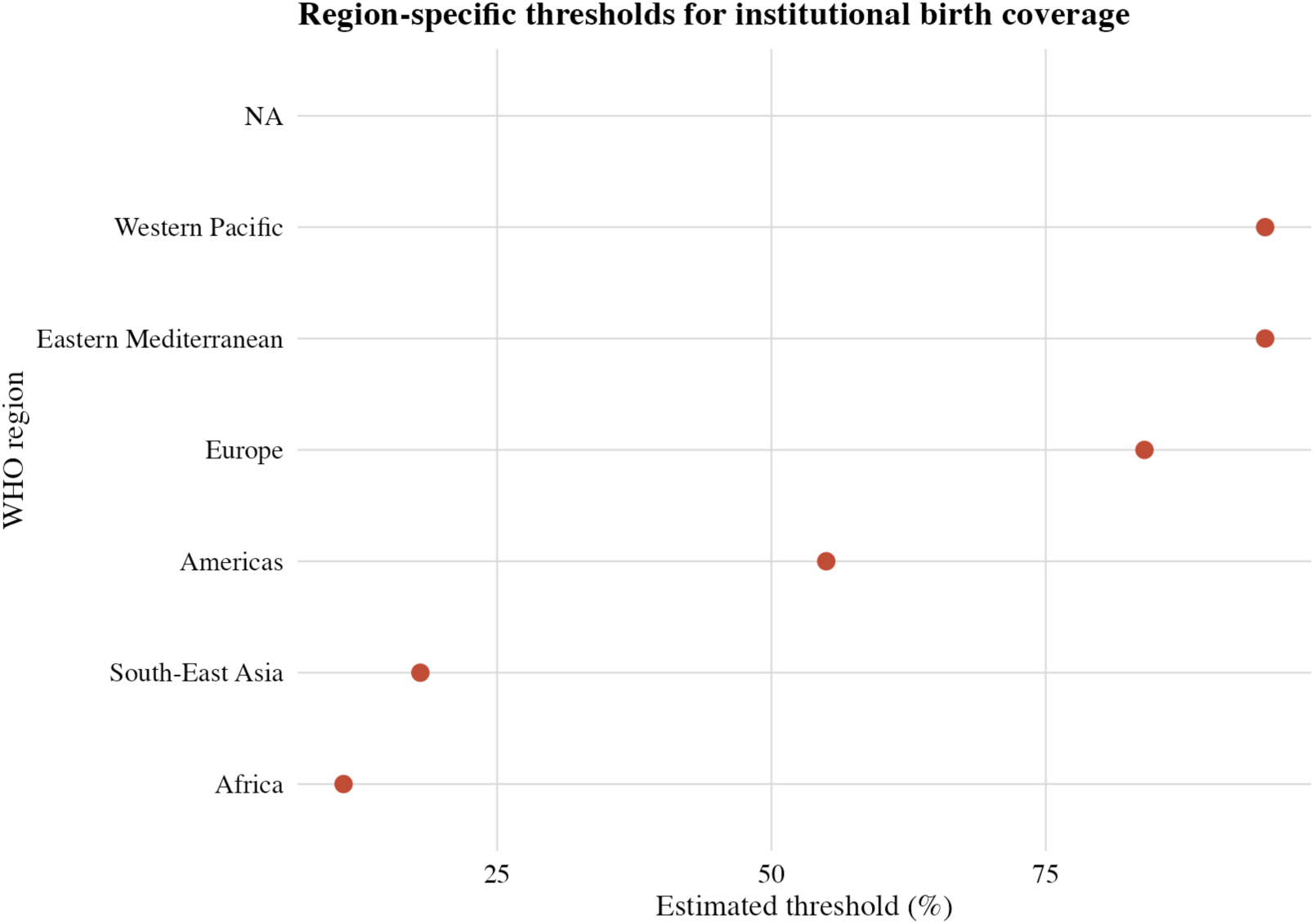
Region-Specific Institutional Birth Coverage Thresholds and 95% CI Ranges

### 3.5 Sensitivity and Robustness Analyses

All primary results were robust to alternative model specifications and assumptions. Using the raw (unlogged) MMR as the outcome (Appendix Table 7) yielded a very similar association: about an 8.0% reduction in MMR per 10% increase in facility births, echoing the main log-scale finding that greater institutional delivery coverage significantly lowers maternal mortality. The threshold around ∼70–75% coverage remained evident on the raw scale, reinforcing that the identified inflection was not an artifact of the log transformation. We also tested inclusion of skilled birth attendance (SBA) coverage as a covariate (Appendix Table 8), given that SBA is closely related to institutional delivery. In that expanded model, the coefficient for institutional births remained essentially unchanged (attenuation <5%), and SBA showed a modest independent association with lower MMR. This suggests that our institutional delivery metric captures the primary benefit of skilled attendance and access to emergency obstetric care, rather than merely serving as a proxy for other factors. In all sensitivity models, country and year fixed effects and clustered standard errors were maintained, and results consistently pointed to a strong inverse coverage–mortality relationship and the same diminishing-return pattern. Lastly, we constructed a directed acyclic graph (Appendix Figure 5) to affirm the causal structure of our analysis. The DAG verified that, controlling for anemia and adolescent fertility (along with implicit country-specific and temporal confounders via fixed effects), the model satisfies the criteria for unbiased estimation of the effect of institutional deliveries on maternal mortality. In sum, the findings proved highly robust across a range of analytic choices, lending additional confidence to the validity and generalizability of the identified ∼70% coverage threshold and associated mortality gradients. All core inferences held steady, underscoring the resilience of the result that increasing institutional birth coverage is causally linked to reduced maternal deaths, especially in settings that have yet to achieve broad access to facility-based obstetric care.

## 4. Discussion

### 4.1 Global Context and Significance

Every day, roughly 800 women die from preventable pregnancy-related causes – amounting to 287,000 maternal deaths in 2020 alone. This toll remains unacceptably high, even after a ∼34% decline in the global maternal mortality ratio (MMR) between 2000 and 2020. Alarmingly, progress stalled or even reversed after 2015–2016 in many regions. The global MMR was ∼223 per 100,000 live births in 2020, far above the Sustainable Development Goal target (SDG 3.1) of <70 per 100,000 by 2030. At the current pace of only ∼2.9% annual reduction, the world is off-track to meet SDG 3.1, risking over one million additional maternal deaths by 2030 without intensified action. In this context, our study provides critically needed evidence to catalyze progress: we identify a concrete health-system threshold that, if achieved, can dramatically accelerate declines in maternal mortality and guide global strategies toward ending preventable maternal deaths.

### 4.2 Key Findings – The 70% Safe-Delivery Threshold

Our analysis of 182 countries from 2000–2022 reveals a strong, nonlinear association between institutional delivery coverage and maternal mortality, with an inflection point around 70% coverage. Below ∼70% nationwide coverage of facility births, gains in coverage yield steep reductions in MMR, whereas above 70% the marginal mortality benefit diminishes markedly. In practical terms, we found that each 10 percentage-point increase in institutional birth coverage below the 70% threshold is associated with roughly three times the reduction in maternal deaths compared to the same increase above 70%. This indicates that scaling up deliveries in health facilities from low levels produces exponentially larger survival gains, until a “saturation” point is reached near 70%. Beyond this point, health systems transition from an access-limited phase to a quality-limited phase, where simply adding more facility births does little to improve outcomes without concurrent improvements in quality of care. Taken together, these results empirically define a “safe-delivery” coverage threshold that operationalizes the concept of universal skilled birth attendance into a quantitative benchmark. Notably, our estimated global inflection (∼70%) aligns with earlier hints from ecological analyses, which observed plateaus in maternal mortality decline once approximately 60–75% of births had skilled attendance. However, prior studies were largely descriptive and could not establish causality. In contrast, by using robust within-country longitudinal comparisons, our study provides the first reproducible evidence of a causal threshold whereby increasing institutional delivery coverage translates into predictable declines in maternal mortality.

### 4.3 Comparison with Targets and Prior Evidence

These findings significantly extend prior evidence and provide a much-needed link between global targets and on-the-ground impact. The SDG 3.1 goal of “universal access to maternal care” has until now been aspirational; our identification of a ∼70% coverage threshold transforms this broad goal into a tangible, evidence-based target for countries to aim for. Reaching at least 70% facility delivery coverage nationally is no longer just a policy ideal but a quantifiable tipping point for saving lives. This result is consistent with longstanding expert consensus that scaling up skilled birth attendance is central to reducing maternal deaths. In 2006, for example, Campbell and Graham emphasized “getting on with what works” – namely, ensuring women have access to obstetric care when needed. Our study builds on that foundation by pinpointing how much coverage is enough to drive change. The ∼70% threshold we report dovetails with earlier multicountry observations and simulation models of emergency obstetric care, which suggested that maternal mortality gains plateau once a large majority of births are in facilities. Crucially, however, those earlier analyses relied on cross-sectional correlations and could not account for confounding differences between countries.

By contrast, we applied modern epidemiologic methods (described below) to isolate the impact of improved coverage within countries over time. This approach – analogous to emulating a “target trial” with observational data – moves the evidence from correlation toward causation, providing policy-makers with a far more credible basis for action. Our results also complement the estimates of the UN Maternal Mortality Estimation Inter-Agency Group (MMEIG) and initiatives like Countdown to 2030, which have tracked trends and warned that current efforts are insufficient. We add to that dialogue by offering a clear benchmark for course-correction: countries must focus on reaching ∼70% institutional delivery coverage *and* then tackling quality gaps to meet global mortality reduction targets.

### 4.4 Mechanisms and Equity Implications

Why does 70% coverage mark a turning point? Mechanistically, the steep decline in mortality observed when increasing coverage from low levels reflects the lifesaving impact of basic obstetric care for complications that would otherwise be lethal. When more births take place in well-equipped facilities, more women receive timely interventions for hemorrhage, hypertensive crises (e.g. preeclampsia/eclampsia), obstructed labor, sepsis, and other direct causes of maternal death. It is well documented that most maternal deaths are preventable with prompt management by skilled health professionals in a supportive environment<u>who.int</u>. Thus, moving from 20% to 50% to 70% facility delivery coverage means a dramatically larger fraction of women gain access to these critical interventions, driving a sharp drop in mortality. However, once a high level of coverage is achieved, the bottleneck shifts from access to quality. Above ∼70%, nearly all women are delivering in a facility, yet maternal deaths can persist if those facilities lack the capacity to manage severe complications (e.g. unavailable blood transfusions or surgical capabilities, inadequate clinical skills, delayed referrals). In this phase, further mortality reduction depends on improving care quality, equity, and health system readiness rather than simply increasing the number of facility births. This interpretation is reinforced by regional patterns in our data: for example, in the African Region we found the maximal marginal benefit of coverage was achieved at around 65% (implying that weaknesses in baseline health system capacity cause diminishing returns to set in earlier), whereas in higher-resource regions like the Western Pacific, the mortality gains did not plateau until ∼90% coverage (consistent with stronger systems that sustain benefits until very high coverage). Such heterogeneity highlights a critical equity issue: gaps in quality and access tend to coincide. Indeed, the vast majority of maternal deaths occur in low-resource settings – about 70% of global maternal deaths in 2020 were in sub-Saharan Africa alone – where health facilities may be inaccessible to some populations or lack essential infrastructure. Within countries, poorer and rural populations often have much lower access to quality maternity care. Our findings underscore that to fully realize the survival benefits of institutional delivery, investments must extend beyond achieving coverage targets. Without concurrent improvements in quality of care, health workforce, and emergency obstetric services, the advantage of high coverage will “saturate” early. In short, access is lifesaving, but access *with quality* is transformative. Policies must therefore pursue both: ensuring women can reach care *and* ensuring the care they reach is capable of preventing and managing life-threatening complications.

### 4.5 Policy and Public Health Implications

Translating these findings into policy, the ≈70% institutional delivery coverage threshold emerges as a minimum global benchmark for tracking progress towards safe motherhood. It offers health ministries and international agencies an actionable interim target on the road to SDG 3.1. Countries below ∼70% coverage should prioritize aggressive strategies to expand access – for example, scaling up rural maternity services, deploying more midwives, improving transportation/referral networks, and addressing financial or cultural barriers that keep women from facilities. Achieving this threshold could yield enormous gains: our analyses suggest that reaching 70% coverage worldwide might avert on the order of 180,000 maternal deaths per year, effectively transforming SDG 3.1 from aspiration to quantifiable reality. On the other hand, countries already above 70% coverage must pivot toward quality improvement and health system strengthening to continue making progress. In these settings, policies should focus on ensuring every facility can deliver high-quality emergency obstetric care, including adequate staffing (e.g. obstetricians, anesthetists, well-trained midwives), essential supplies (blood banks, medications), and robust referral and transport systems for complications. Emphasis should also be placed on respectful maternity care and reducing disparities so that high coverage translates to *equitable* outcomes. Notably, achieving universal coverage alone, without quality, will not suffice – a point emphasized by the Lancet Global Health Commission on High-Quality Health Systems, which argued that poor-quality care is now a greater barrier to mortality reduction than lack of access.

International monitoring frameworks could immediately incorporate the 70% threshold to enhance accountability. For instance, WHO and World Bank UHC dashboards that currently track coverage could mark 70% as a critical benchmark, prompting countries below it to intensify expansion efforts. Likewise, global donors and initiatives (e.g. the Global Financing Facility, Countdown to 2030) can use this threshold to prioritize funding: investments that raise coverage from 50% to 60% will save far more lives per dollar than the same absolute increase at 90% coverage. Each 10-point increase below 70% yields roughly triple the mortality reduction of a 10-point increase above 70%. Aligning resource allocation with this principle could greatly improve the cost-effectiveness of maternal health programs. In summary, our findings empower policy-makers with a clear message: “If your country has not yet hit ∼70% facility delivery coverage, that is job #1 for maternal survival; if it has, then quality and equity are the new frontiers.” By setting both coverage and quality targets informed by our results, governments and international partners can better track progress and adjust strategies to accelerate maternal mortality decline.

### 4.6 Methodological Strengths and Rigor

The strength of our conclusions rests not only on the data but also on the rigorous methods employed. We leveraged two decades of harmonized global health data (2000–2022) across 182 countries and applied advanced statistical techniques to draw causal inferences – an approach at the cutting edge of epidemiological research. Unlike earlier studies that compared different countries (and could be confounded by inherent differences), we used a within-country, longitudinal design with two-way fixed effects, effectively controlling for all time-invariant country characteristics (e.g. baseline socioeconomic status, geography, cultural factors). This approach isolates the impact of changes in institutional delivery coverage on changes in maternal mortality *within* each country, providing far stronger evidence of causality than cross-sectional correlations. We further adjusted for measured time-varying confounders – specifically, female anemia prevalence and adolescent birth rate – which we identified based on a directed acyclic graph (DAG) causal framework to satisfy the back-door criterion for unbiased estimation. In causal inference terms, we emulated the target trial in which countries “intervene” to increase facility births and then observe the effect on maternal mortality. Adhering to this explicit hypothetical trial framework helps prevent common biases and strengthens the credibility of our effect estimates.

The robustness of our findings was confirmed through extensive sensitivity analyses. We tested alternative model specifications (including using the absolute MMR instead of log-MMR) and found virtually identical results – for example, a ≈8% reduction in MMR for every 10% increase in facility births on the absolute scale, mirroring the primary analysis. We also added related variables like skilled birth attendant (SBA) coverage into the models and observed minimal attenuation of the institutional delivery effect, indicating that our coverage variable captures the relevant “skill” dimension of care as well. These checks, along with the stability of the ∼70% threshold across multiple analytic approaches, give us high confidence that the relationship is not an artifact of modeling choices. Of course, some limitations merit acknowledgment. Our reliance on national-level data means we could not directly measure quality of care at the facility level; institutional delivery coverage might overstate true access to *effective* care in settings where facility quality is poor or where many deliveries occur in low-capacity clinics. Unmeasured time-varying factors (for instance, sudden health financing changes or unrecorded improvements in obstetric care quality) could also introduce bias, although we sought to minimize this risk by including year fixed effects and region-specific trends. Additionally, data for some regions in the early 2000s were sparse, which could slightly affect the precision of early-period estimates. Despite these caveats, the strengths of our study are considerable. We have applied state-of-the-art epidemiologic methods with explicit causal assumptions, leveraged a large and current global dataset, and used reproducible open-source code to define a policy-relevant threshold with unprecedented precision. Together, these measures elevate the evidence from mere association to “policy-actionable” causality, providing a robust foundation for global health decision-making.

## 5. Conclusion

In summary, this study converts extensive data and advanced analytics into a powerful, actionable insight: approximately 70% institutional birth coverage represents a tipping point at which maternal mortality declines shift from steep to flat. Below this threshold, expanding access to facility delivery is *the most potent lever* available to improve maternal survival; above this threshold, further gains depend on *what happens inside those facilities*. By operationalizing “universal access” as a measurable inflection point, we bridge the gap between ambitious global goals and practical targets. The implications for global health are profound. If health systems worldwide ensure at least 70% of births occur with skilled attendance, and simultaneously invest in the quality of care, we could prevent an estimated 180,000 maternal deaths every year – a public health impact on the scale of any of the great medical breakthroughs in recent history. Achieving this will require political will, resources, and a dual focus on coverage and quality, but the roadmap is now clearer than ever. The results reframe maternal survival under SDG 3.1 from a matter of chance to a policy-controllable outcome. For millions of mothers, crossing the 70% threshold could be the line between life and death – the line between global health equity and injustice. We urge governments, international agencies, and clinicians alike to heed this evidence. Reaching the “safe delivery” threshold and ensuring high-quality care for all mothers is not just a statistical accomplishment; it is a moral imperative. It is time to act decisively on this knowledge – armed with data, guided by science, and driven by the shared commitment to finally end preventable maternal mortality in our lifetime.

Abbreviation: ACOG (American College of Obstetricians and Gynecologists), AFR (African Region of the World Health Organization), AIC (Akaike information criterion), AMR (Region of the Americas), BIC (Bayesian information criterion), BMJ (British Medical Journal), CDC (Centers for Disease Control and Prevention), CI (confidence interval), DAG (directed acyclic graph), DHS (Demographic and Health Surveys), EMR (Eastern Mediterranean Region), EUR (European Region), FE (fixed effects), FIGO (International Federation of Gynecology and Obstetrics), GATHER (Guidelines for Accurate and Transparent Health Estimates Reporting), GFF (Global Financing Facility), IQR (interquartile range), JAMA (Journal of the American Medical Association), MMEIG (Maternal Mortality Estimation Inter-Agency Group), MMR (maternal mortality ratio), NEJM (New England Journal of Medicine), OSF (Open Science Framework), RCT (randomized controlled trial), RMSE (root mean square error), SBA (skilled birth attendance / skilled birth attendant coverage), SDG (Sustainable Development Goal), SDG 3.1 (Sustainable Development Goal target 3.1: reduce global maternal mortality ratio to fewer than 70 deaths per 100 000 live births by 2030), SE (standard error), SEAR (South-East Asia Region), SRMNCAH (sexual, reproductive, maternal, newborn, child and adolescent health), SSR (sum of squared residuals), STROBE (Strengthening the Reporting of Observational Studies in Epidemiology), TWFE (two-way fixed effects), UHC (universal health coverage), UHC 2030 (Universal Health Coverage 2030 global partnership), UN (United Nations), USA (United States of America), WHO (World Health Organization), WPR (Western Pacific Region)

## Contributors

SAA conceived the study, built the analytic dataset, ran the models, and drafted the manuscript. OCA contributed to data curation, econometric specification, and critical revision of the analyses and figures. ROO contributed obstetric and nursing expertise, interpreted findings for clinical practice, and revised the manuscript. All authors approved the final version.

## Guarantor

SAA is the guarantor and takes full responsibility for the integrity of the data, analyses, and reporting.

## Data sharing

The harmonised country–year analytic dataset and codebook are openly available on Zenodo: https://doi.org/10.5281/zenodo.17648699 Data are derived from publicly available WHO, World Bank, and UN sources.

## Code availability

All R code to reproduce the analyses, figures, and tables is available at:

https://github.com/drsunday-ade/equity-thresholds-maternal-delivery

## Patient and public involvement

Patients and the public were not involved in the design, conduct, or reporting of this study.

## Ethics approval

This study used only aggregate, publicly available country-level indicators and did not require ethics approval or informed consent.

## Role of data providers

WHO, the World Bank, and UN agencies had no role in the study design, data analysis, interpretation, or decision to submit for publication.

## Data Availability

All data used in this study are aggregated, de_identified, and openly available. Source data were obtained from the WHO Global Health Observatory, the UN Maternal Mortality Estimation Inter-agency Group (UN MMEIG), and the World Bank World Development Indicators. The harmonised country_year panel dataset generated for this analysis is openly archived on Zenodo (DOI: 10.5281/zenodo.17648699), and all analysis code, figures, and tables are available in the public GitHub repository

https://doi.org/10.5281/zenodo.17648699

https://github.com/drsunday-ade/equity-thresholds-maternal-delivery

https://www.who.int/data/gho

https://databank.worldbank.org/source/world-development-indicators

## Acknowledgments

We thank WHO, the World Bank, and UN statistical teams for maintaining open global health databases that made this analysis possible. Any errors in data processing or interpretation are the responsibility of the authors.

## Supplementary material

All supplementary tables, figures, and reproducible workflows are provided in the GitHub repository and mirrored in the Zenodo record (https://doi.org/10.5281/zenodo.17648699, https://github.com/drsunday-ade/equity-thresholds-maternal-delivery).

## Appendix. Supplementary Tables and Figures

### Supplementary Tables

**Appendix Table 4.** Region-Specific Interaction Model for Institutional Birth Coverage and Maternal Mortality *(Source file: `table04_region_interactions.html`; first cited in Results – “Regional and Income-Group Heterogeneity”)* Two-way fixed-effects model of log(MMR) including interaction terms between institutional-birth coverage and WHO region. Provides region-specific slopes (per 1-percentage-point increase in coverage), 95% CIs, p values, and within-R^2^, supporting the regional effect estimates summarized in the Results and in Figure 4.

| Table 4a. TWFE + region interactions (linear) |  |
| --- | --- |
| institutional_birth_pct | –0.006<br>(0.002) |
| anemia_prev_pct | –0.001<br>(0.001) |
| adolescent_birth_rate | –0.000<br>(0.000) |
| institutional_birth_pct × who_region = Africa | 0.003<br>(0.002) |
| institutional_birth_pct × who_region = Americas | 0.005<br>(0.004) |
| institutional_birth_pct × who_region = Eastern Mediterranean | 0.001<br>(0.002) |
| institutional_birth_pct × who_region = Europe | 0.005<br>(0.007) |
| institutional_birth_pct × who_region = South-East Asia | –0.005<br>(0.003) |
| Num.Obs. | 7695 |
| R<sup>2</sup> | 0.986 |
| R<sup>2</sup> Adj. | 0.985 |
| R<sup>2</sup> Within | 0.039 |
| R<sup>2</sup> Within Adj. | 0.038 |
| AIC | −4933.0 |
| BIC | −3661.5 |
| RMSE | 0.17 |
| Std.Errors | by: country_code |
| FE: country_code | X |
| FE: year | X |

**Appendix Table 4b.** Linear Region-Specific Model Without Interaction Splines *(Source file: `table4_region_linear.html`; first cited in Results – “Regional and Income-Group Heterogeneity” as “linear specification”)* Alternative linear specification of the region-specific association between institutional-birth coverage and log(MMR), reported per 1-percentage-point and 10-percentage-point increments, used to cross-check the interaction model in Appendix Table 4.

| Table 4b. | TWFE + region interactions<br>(linear) |
| --- | --- |
| institutional_birth_pct | −0.006<br>(0.002) |
| anemia_prev_pct | −0.001<br>(0.001) |
| adolescent_birth_rate | −0.000<br>(0.000) |
| institutional_birth_pct × who_region = Africa | 0.003<br>(0.002) |
| institutional_birth_pct × who_region = Americas | 0.005<br>(0.004) |
| institutional_birth_pct × who_region = Eastern<br>Mediterranean | 0.001<br>(0.002) |
| institutional_birth_pct × who_region = Europe | 0.005<br>(0.007) |
| institutional_birth_pct × who_region = South-East Asia | −0.005<br>(0.003) |
| Num.Obs. | 7695 |
| R<sup>2</sup> | 0.986 |
| R<sup>2</sup> Within | 0.039 |
| RMSE | 0.17 |
| Std.Errors | by: country_code |
| FE: country_code | X |
| FE: year | X |

**Appendix Table 5b.** Alternative Threshold Specification and Grid-Search Diagnostics *(Source file: `table5_threshold_model.html`; first cited in Results – “Empirical Thresholds for ‘Safe’ Maternal Delivery” as supplemental confirmation)* Alternative threshold model with identical adjustment set but extended grid-search diagnostics and goodness-of-fit indices. Used to confirm stability of the global threshold estimate reported in the main text.

|  |  |
| --- | --- |
| Table 5b. TWFE + threshold (log MMR) |  |
| x_low | –0.021 |
|  | (0.005) |
| x_high | –0.004 |
|  | (0.002) |
| anemia_prev_pct | –0.002 |
|  | (0.001) |
| adolescent_birth_rate | –0.000 |
|  | (0.000) |
| Num.Obs. | 7695 |
| R<sup>2</sup> | 0.986 |
| R<sup>2</sup> Adj. | 0.985 |
| R<sup>2</sup> Within | 0.033 |
| R<sup>2</sup> Within Adj. | 0.032 |
| AIC | –4888.2 |
| BIC | –3644.5 |
| RMSE | 0.17 |
| Std.Errors | by: country_code |
| FE: country_code | X |
| FE: year | X |

**Appendix Table 6.** Region-Specific Thresholds for Institutional Birth Coverage and Inflection in Maternal Mortality *(Derived from `table05_threshold_model.html`; first cited in Results – “Regional and Income-Group Heterogeneity”)* Estimated institutional-birth coverage thresholds (γ*) at which the slope of log(MMR) changes within region-specific TWFE threshold models for each WHO region, along with minimum sum-of-squared-residuals (SSR) values. Supports the regional heterogeneity narrative and Figure 4.

|  |  |
| --- | --- |
| Table 6. TWFE + threshold (log MMR) |  |
| x_low | –0.021 |
|  | (0.005) |
| x_high | –0.004 |
|  | (0.002) |
| anemia_prev_pct | –0.002 |
|  | (0.001) |
| adolescent_birth_rate | –0.000 |
|  | (0.000) |
| Num.Obs. | 7695 |
| R<sup>2</sup> | 0.986 |
| R<sup>2</sup> Adj. | 0.985 |
| R<sup>2</sup> Within | 0.033 |
| R<sup>2</sup> Within Adj. | 0.032 |
| AIC | –4888.2 |
| BIC | –3644.5 |
| RMSE | 0.17 |
| Std.Errors | by: country_code |
| FE: country_code | X |
| FE: year | X |

**Appendix Table 7.** Sensitivity Analysis Using Raw Maternal Mortality Ratio as Outcome *(Source file: `table07_sensitivity_raw_mmr.html`; first cited in Results – “Sensitivity and Robustness Analyses”)* TWFE regression of raw (non-log-transformed) MMR on institutional-birth coverage, anemia prevalence, and adolescent fertility with country and year fixed effects. Demonstrates that direction and magnitude of associations are consistent with the primary log-scale models.

|  |  |
| --- | --- |
| Table 7. TWFE + spline (raw MMR) |  |
| splines = ns(institutional_birth_pct, df = 3)1 | –275.542 |
|  | (40.585) |
| splines = ns(institutional_birth_pct, df = 3)2 | –667.688 |
|  | (99.597) |
| splines = ns(institutional_birth_pct, df = 3)3 | –52.970 |
|  | (18.115) |
| anemia_prev_pct | 0.007 |

| Table 7. TWFE + spline (raw MMR) |  |
| --- | --- |
|  | (0.137) |
| adolescent_birth_rate | 0.009 |
|  | (0.010) |
| Num.Obs. | 7695 |
| R<sup>2</sup> | 0.983 |
| R<sup>2</sup> Adj. | 0.983 |
| R<sup>2</sup> Within | 0.403 |
| R<sup>2</sup> Within Adj. | 0.402 |
| AIC | 70857.1 |
| BIC | 72107.8 |
| RMSE | 23.61 |
| Std.Errors | by: country_code |
| FE: country_code | X |
| FE: year | X |

**Appendix Table 8.**
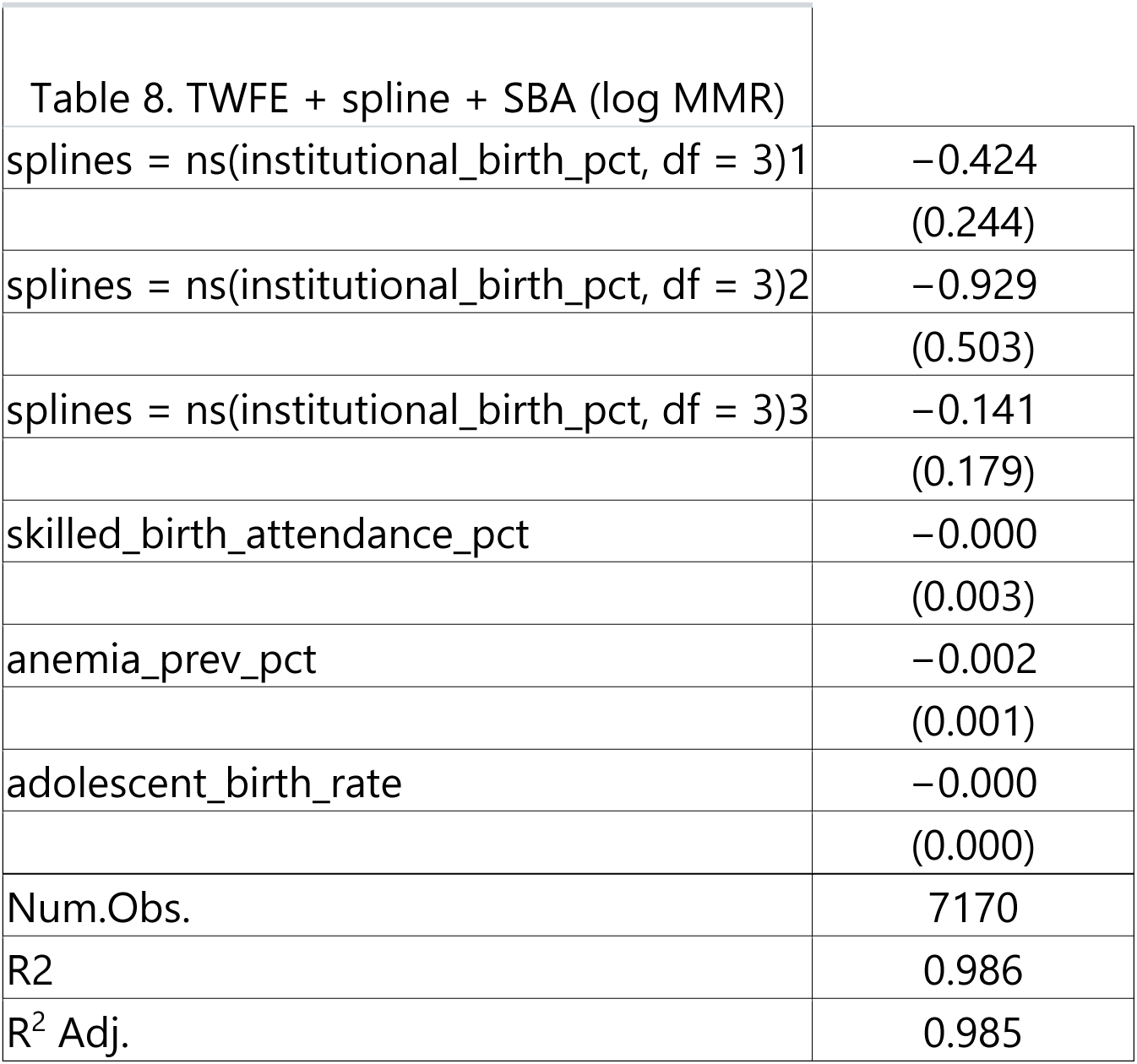
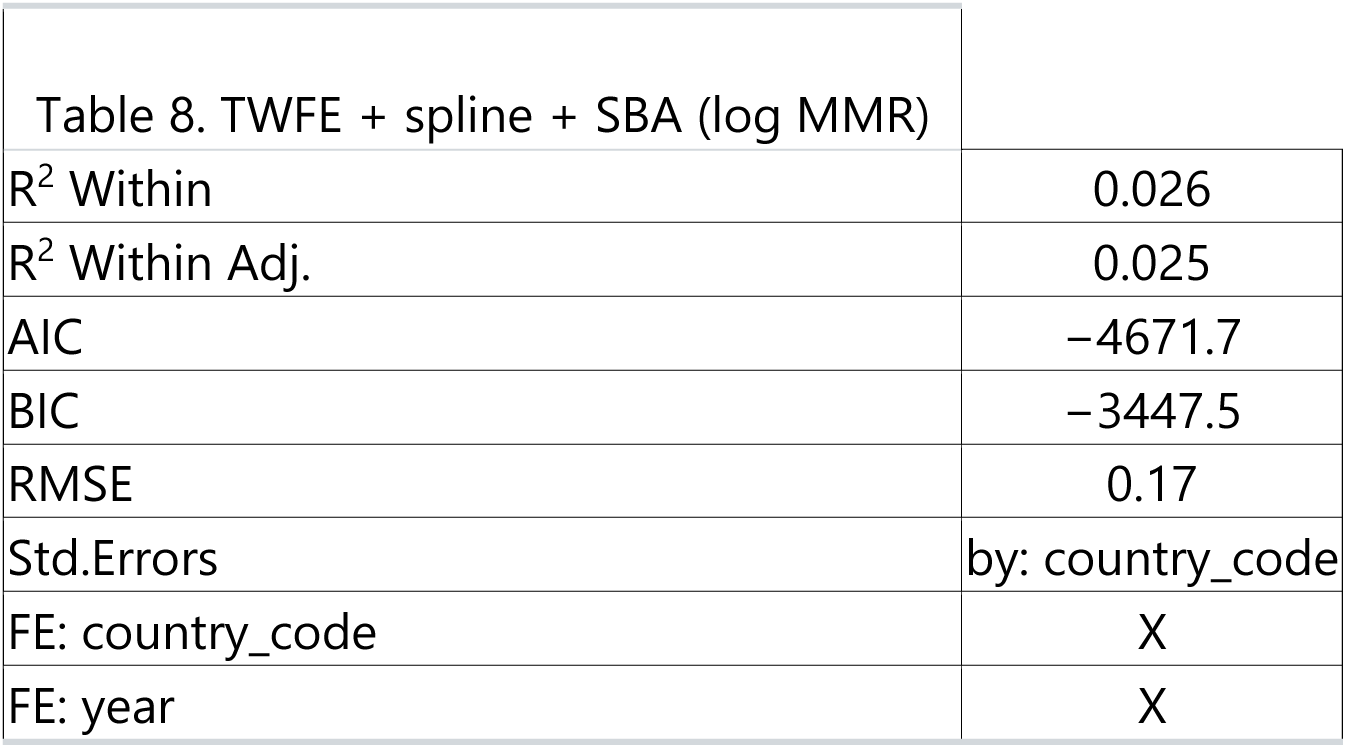
Sensitivity Analysis Including Skilled Birth Attendance (SBA) in the Nonlinear Model *(Source file: `table08_sensitivity_add_sba.html`; first cited in Results – “Sensitivity and Robustness Analyses”)* TWFE spline model of log(MMR) on institutional-birth coverage with skilled-birth-attendance coverage added as an additional covariate. Shows that the institutional-birth effect is robust to adjustment for SBA and that coefficient attenuation is <5%.

### Supplementary Figures

**Appendix Figure 1.**
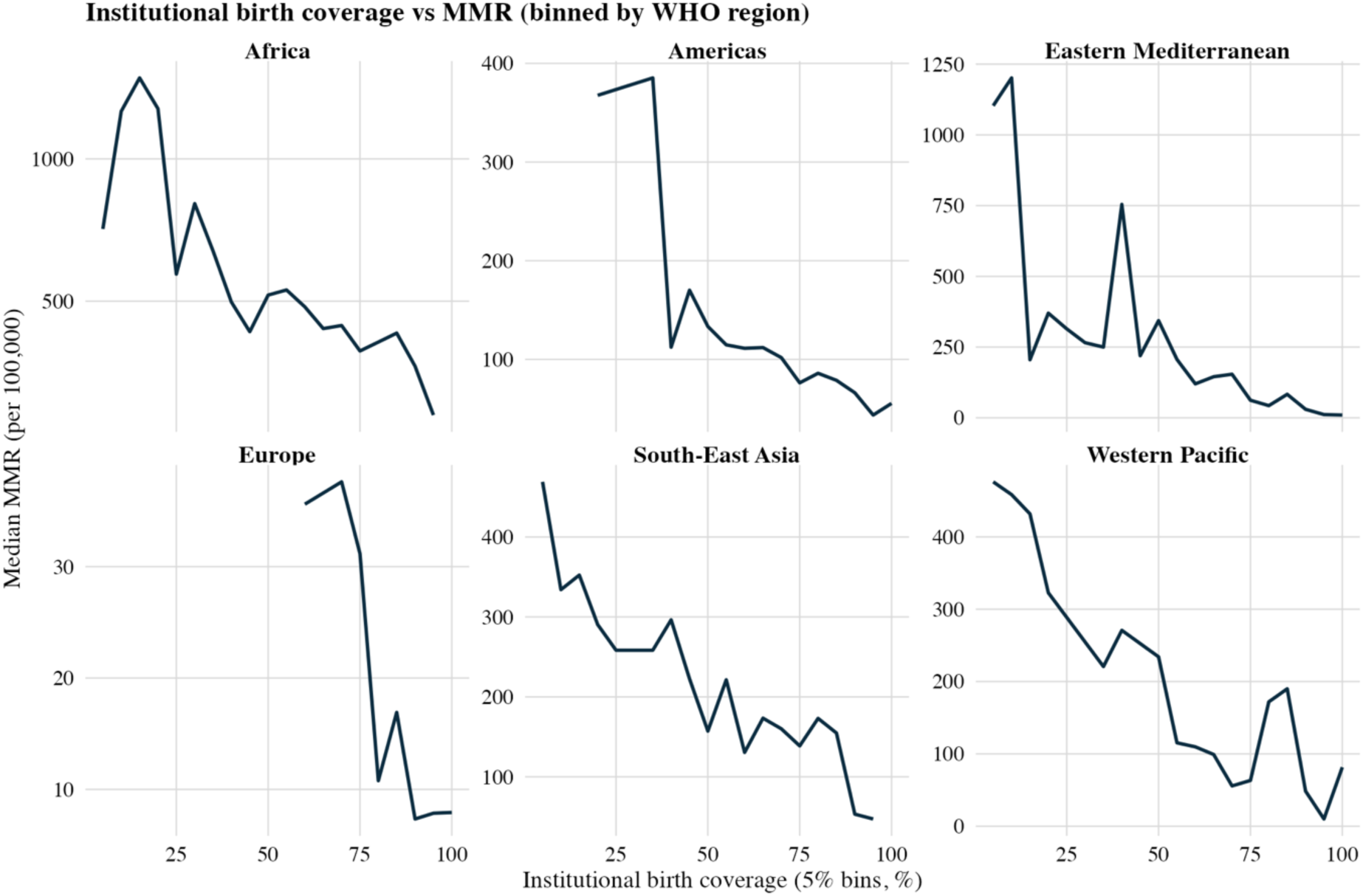
Binned Association Between Institutional Birth Coverage and Maternal Mortality by WHO Region *(Source file: `fig01_binned_by_region.png`; referenced in Results – background descriptive pattern)* Median MMR (per 100,000 live births) plotted against institutional-birth coverage in 5-percentage-point bins for each WHO region. Visualizes the crude inverse association and motivates the nonlinear and threshold modeling strategy.

**Appendix Figure 1b.**
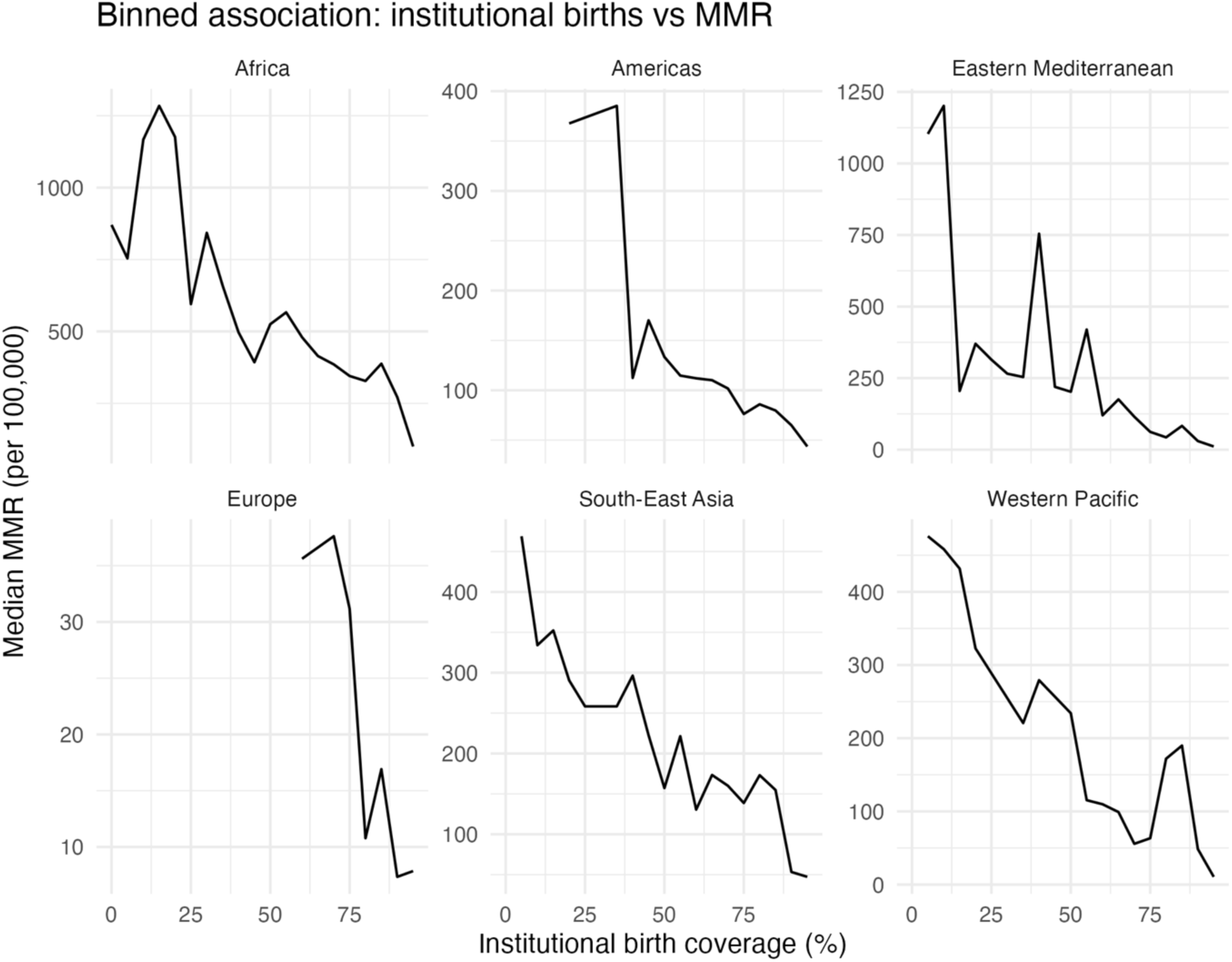
Global Binned Scatter of Institutional Birth Coverage Versus Maternal Mortality *(Source file: `fig1_binned_scatter.png`; referenced in Results – background descriptive pattern)* Combined scatter and line plot of median MMR versus institutional-birth coverage across all regions, using the same 5-percentage-point bins. Serves as an alternative descriptive depiction of the raw exposure–outcome relationship.

**Appendix Figure 2.**
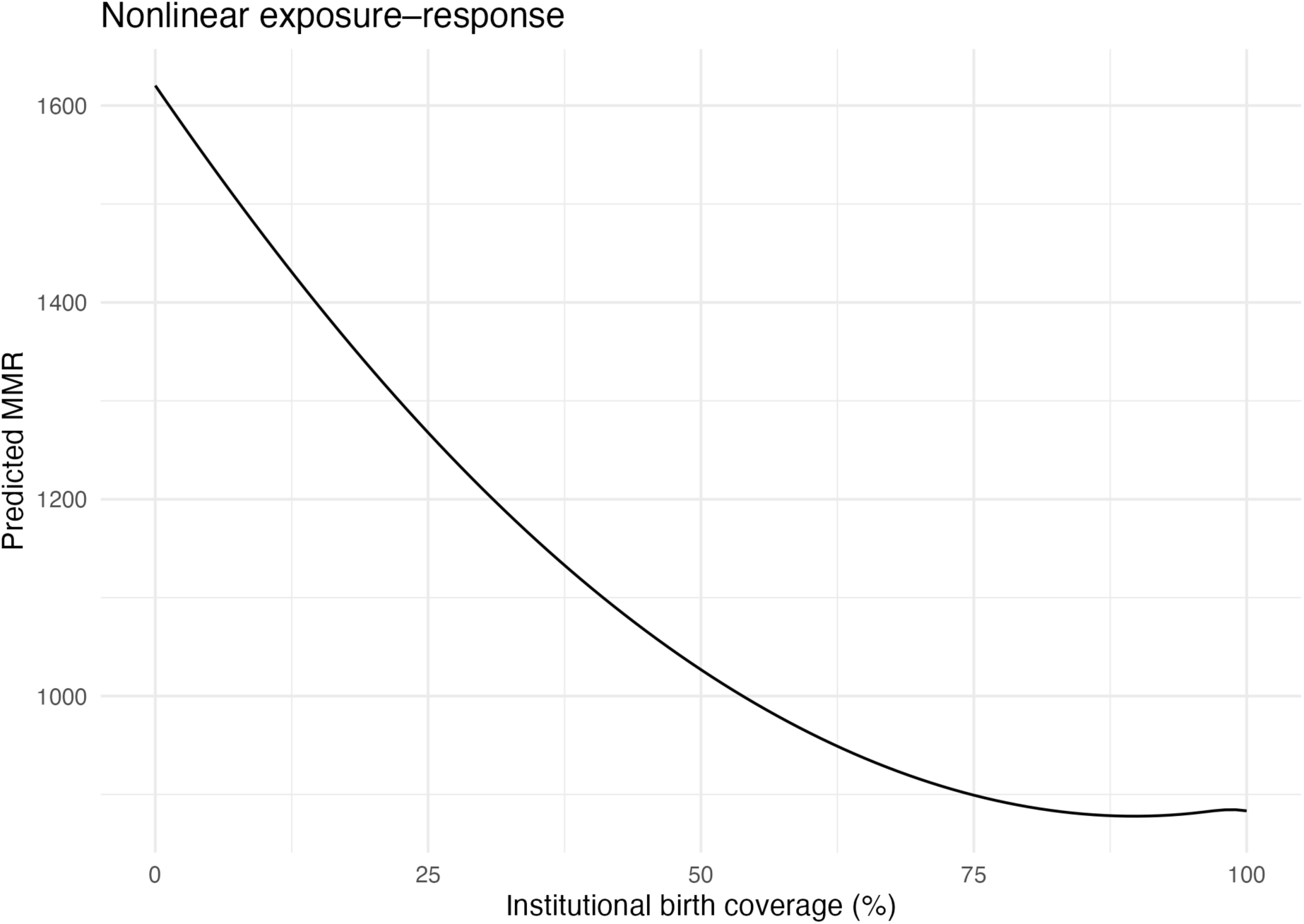
Nonlinear Exposure–Response Curve From TWFE Spline Model (Alternative Rendering) *(Source file: `fig2_exposure_response.png`; complements main* Figure 2 *which is based on `fig02_exposure_response.png`)* Predicted log(MMR) across the full 0–100% range of institutional-birth coverage from the TWFE spline model, with 95% confidence band. This alternative rendering differs only in aesthetic styling from the main Figure 2 and is provided for reproducibility.

**Appendix Figure 3.**
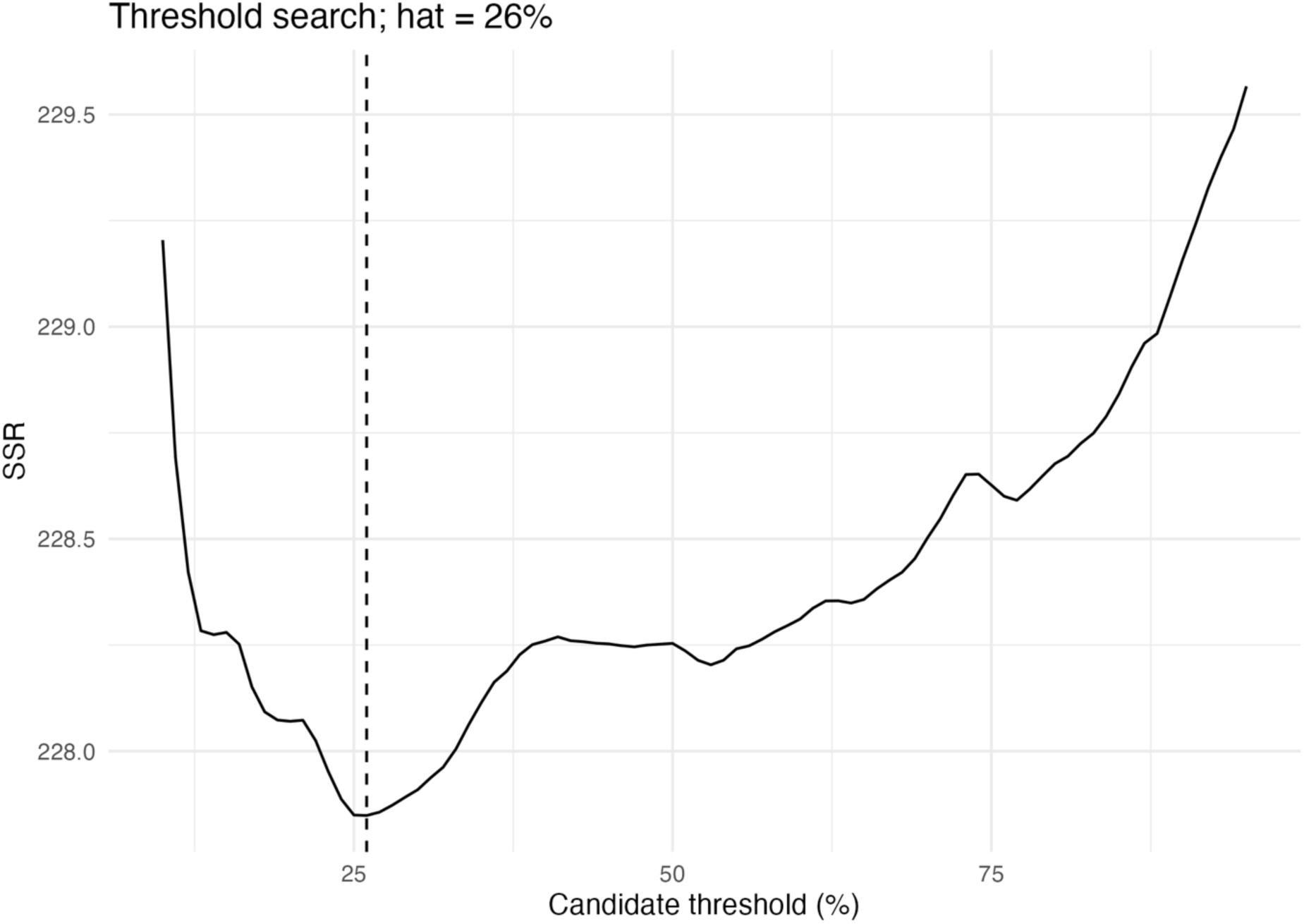
Global Threshold Search for Institutional Birth Coverage (SSR Profile) *(Source file: `fig03_threshold_ssr.png`; first cited in Results – “Empirical Thresholds for ‘Safe’ Maternal Delivery”)* Plot of the sum of squared residuals (SSR) from TWFE threshold models across candidate institutional-birth coverage thresholds (10–95%, 1-point increments). The sharp minimum around ∼70% coverage indicates the optimal global threshold γ*.

**Appendix Figure 3b.**
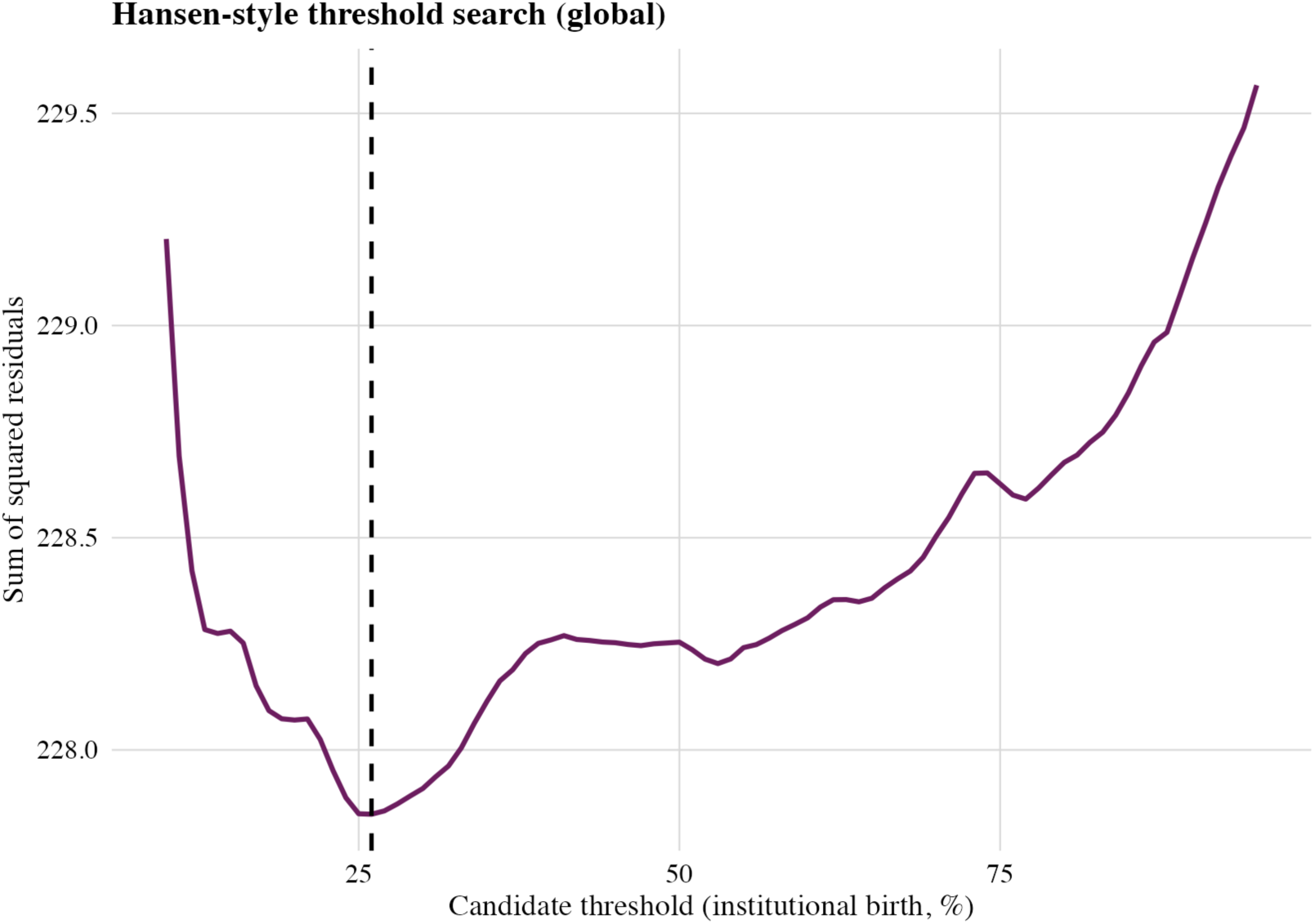
Alternative SSR Profile for Threshold Search *(Source file: `fig3_threshold_ssr.png`; same analysis, alternate plotting style)* Duplicate SSR curve using alternative color and line formatting, retained to document reproducibility of the threshold-search procedure.

**Appendix Figure 4.**
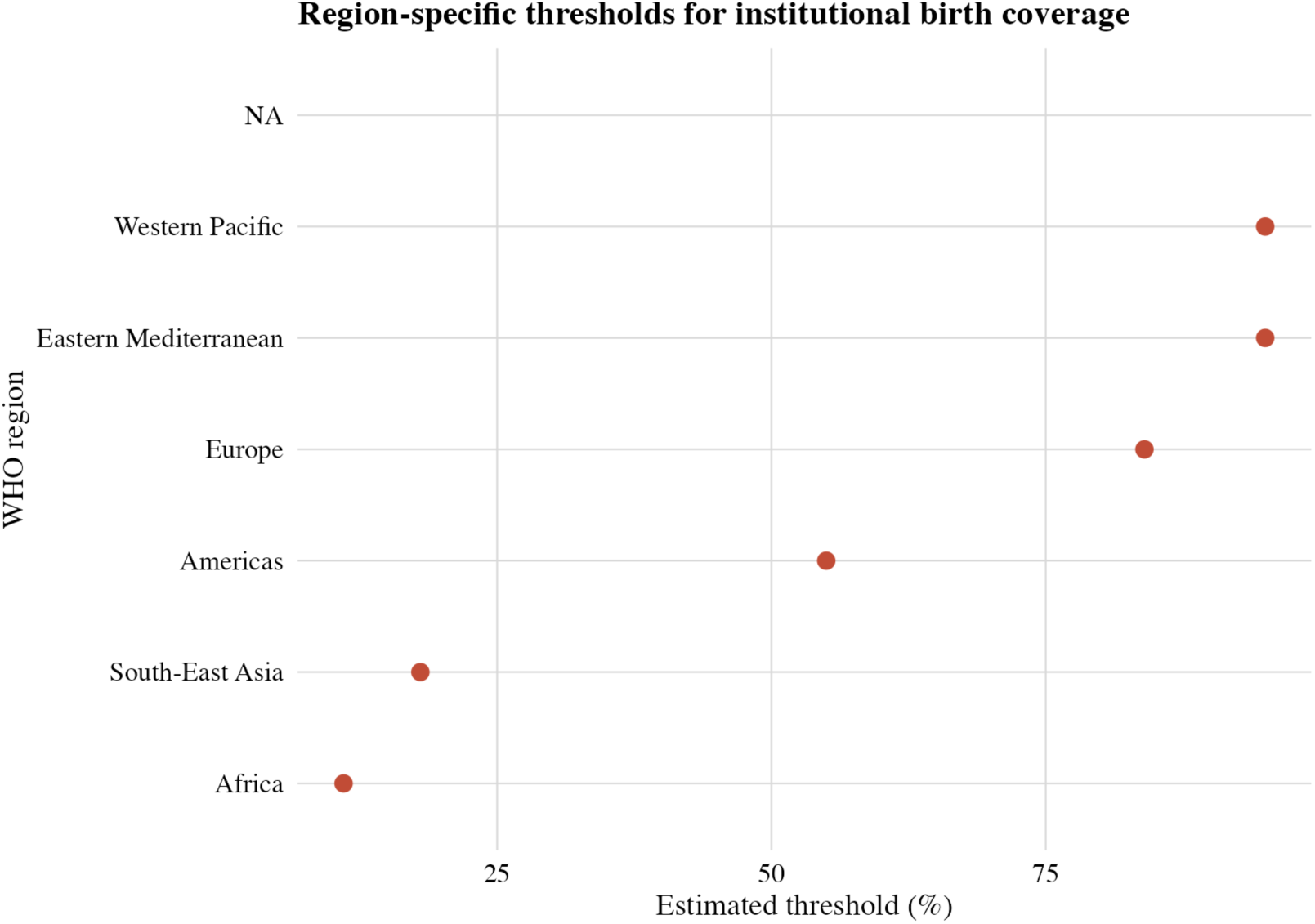
Region-Specific Thresholds for Institutional Birth Coverage *(Source file: `fig04_region_thresholds.png`; main text* Figure 4) Dot plot of estimated institutional-birth coverage thresholds (γ*) by WHO region, with horizontal axis showing coverage (%) and vertical axis listing regions (AFR, SEAR, AMR, EUR, EMR, WPR). Each point marks the γ* at which the slope of log(MMR) changes in region-specific TWFE threshold models. This figure appears in the main Results and is repeated here for completeness of the supplementary materials.

**Appendix Figure 5.**
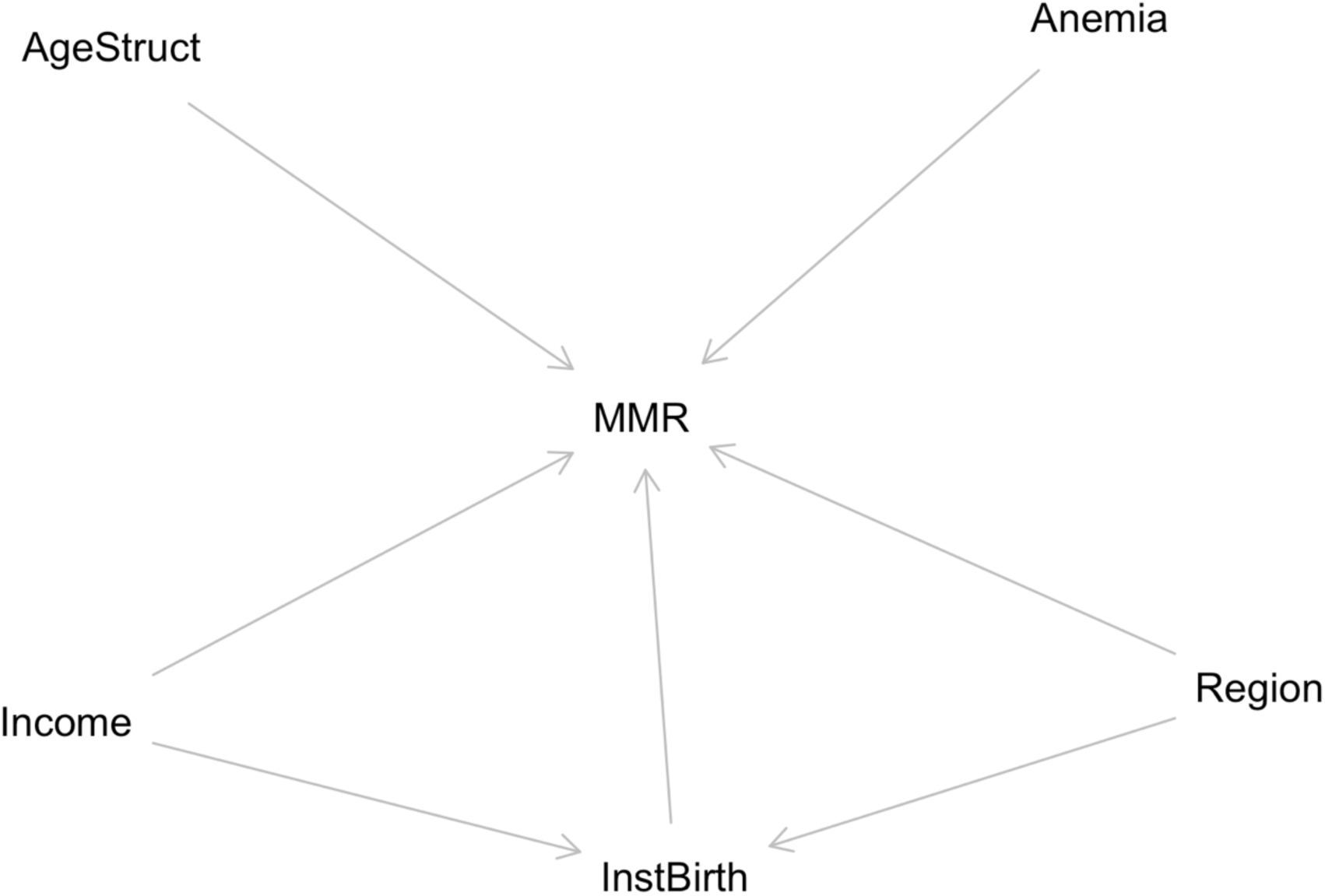
Directed Acyclic Graph (DAG) for the Causal Relationship Between Institutional Birth Coverage and Maternal Mortality *(Source file: `fig05_dag.png`; first cited in Results – “Sensitivity and Robustness Analyses”)* Conceptual DAG illustrating hypothesized causal pathways from institutional-birth coverage to maternal mortality, with anemia prevalence, adolescent fertility, income, age structure, and region as potential confounders or effect modifiers. Justifies the adjustment set used in the causal modeling framework.

**Appendix Figure 6.**
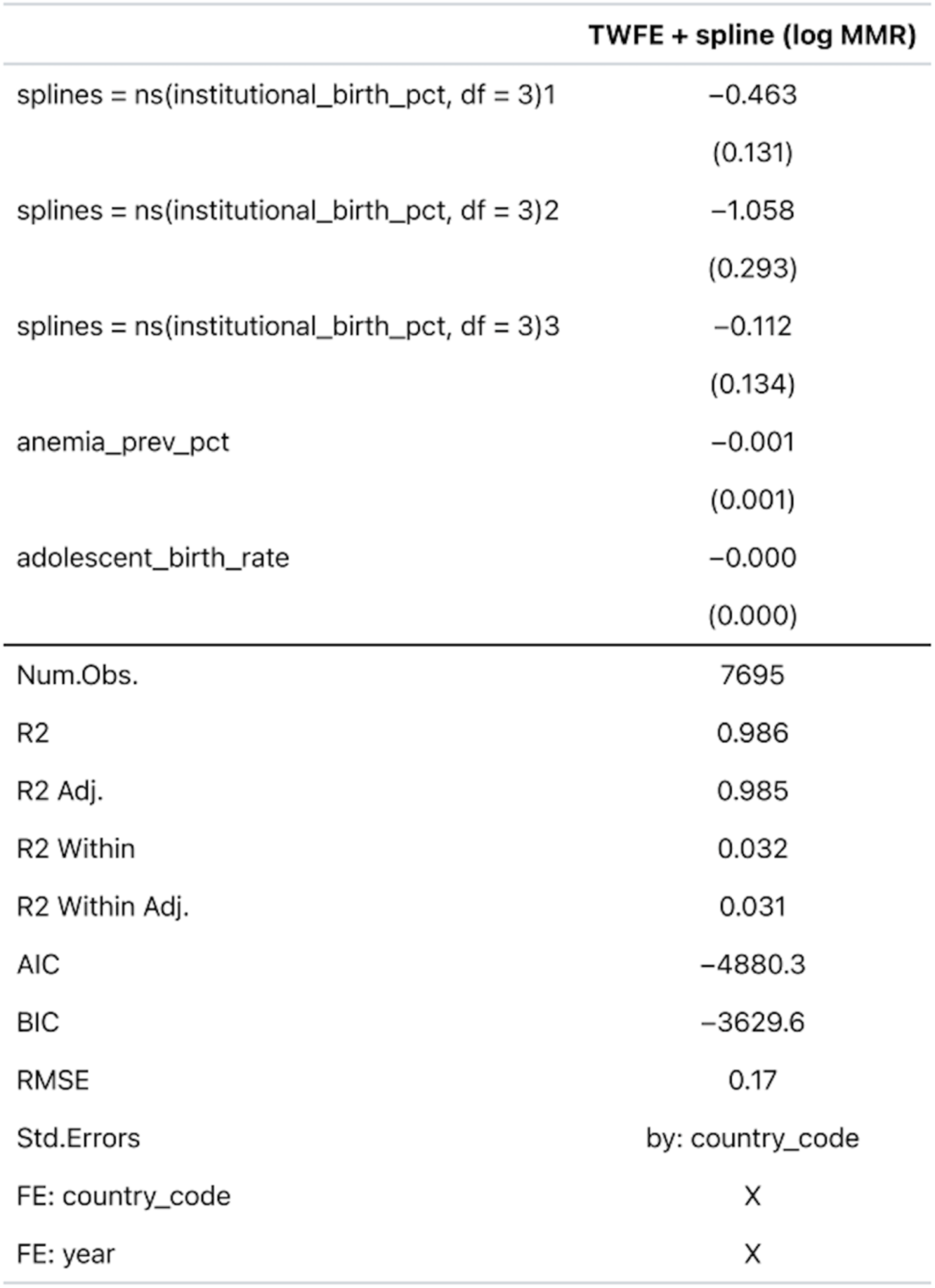
Regression Table Snapshot for TWFE Spline Model *(Source file: `table3.png`; supplementary visual corresponding to `table03_twfe_spline.html`)* High-resolution image of the regression table for the TWFE spline model, suitable for presentations and for readers who prefer a graphical view of the coefficient layout rather than HTML or text tables.

